# The influence of model structure and geographic specificity on predictive accuracy among European COVID-19 forecasts

**DOI:** 10.1101/2025.04.10.25325611

**Authors:** Katharine Sherratt, Rok Grah, Bastian Prasse, Friederike Becker, Jamie McLean, Sam Abbott, Sebastian Funk

## Abstract

Modellers take many approaches to predicting the course of infectious diseases, and achieve a wide range of accuracy in resulting forecasting performance. For example, forecasters vary in their use of different underlying model structures, and in the extent to which they adapt a model to the specific forecast target. However, it has been difficult to evaluate the impact of these choices on subsequent forecast performance. Such evaluations need a comparable sample of forecasting models, while also accounting for varying predictive difficulty among multiple forecast targets. Here, we develop a model-based approach to start addressing these challenges. We apply this to a multi-country multi-model forecasting effort conducted during the COVID-19 pandemic, in order to assess the influence of models’ structure and specificity to the epidemic target on forecast accuracy.

We evaluated 181,851 probabilistic predictions from 47 forecasting models participating in the European COVID-19 Forecast Hub between 8 March 2021 and 10 March 2023, classified by model structure (agent-based, mechanistic, semi-mechanistic, statistical, other); and specificity (the model produced forecasts for either one or multiple locations). We assessed performance of COVID-19 case and death forecasts, measured as the weighted interval score after log-transforming both forecasts and observations. We summarised performance descriptively and compared this to estimates from a generalised additive mixed effects model. We included adjustment for variation between countries over time, the epidemiological situation, the forecast horizon, and among models.

Whilst unadjusted estimates pointed to differences in predictive performance between model structures, after adjustment there was little systematic difference in average performance. Models forecasting for only a single geographic target outperformed those that made predictions for multiple targets, although this was a weak signal. We noted substantial variation in model performance that our approach did not account for.

Understanding the reasons behind forecast performance is useful for prioritising and interpreting modelling work. We showed that valid comparisons of forecast performance depend on appropriately adjusting for the general predictive difficulty of the target. This work was limited by a small sample size of independent models and likely incomplete adjustment for interactions and confounders influencing predictive difficulty. We recommend that multi-model comparisons encourage and document their methodological diversity to enable future studies of underlying factors driving predictive performance.

**Author summary:** Accurately predicting the spread of infectious disease is essential to supporting public health during outbreaks. However, comparing the accuracy of different forecasting models is challenging. Existing evaluations struggle to isolate the impact of model design choices (like model structure or specificity to the forecast target) from the inherent difficulty of predicting complex outbreak dynamics. Our research introduces a novel approach to address this by systematically adjusting for common factors affecting epidemiological forecasts, accounting for multi-layered and non-linear effects on predictive difficulty. We applied this approach to a large dataset of forecasts from 47 different models submitted to the European COVID-19 Forecast Hub. We adjusted for variation across epidemic dynamics, forecast horizon, location, time, and model-specific effects. This allowed us to isolate the impact of model structure and geographic specificity on predictive performance. Our findings suggest that after adjustment, apparent differences in performance between model structures became minimal, while models that were specific to a single location showed a slight performance advantage over multi-location models. Our work highlights the importance of considering predictive difficulty when evaluating across forecasting models, and provides a framework for more robust evaluations of infectious disease predictions.

## Background

The predictive accuracy of infectious disease forecasts varies in time, space, and between forecasting models. When building models for prediction, forecasters have the choice of a range of underlying model structures, with complementary strengths and weaknesses and varying ability to tune a particular model to the forecasting target (1–3). However, it has been difficult to evaluate the impact of these choices on resulting differences in accurate performance. A better understanding would offer insights into the utility of different models, supporting both longer-term model development and short-term selection for accurate prediction during outbreak responses.

A fundamental difference among forecasting models is in the modeller’s choice of underlying structure. This can range on a spectrum of statistical, to mechanistic, reflecting the level of prior understanding and belief in how transmission unfolds in the included population. While statistical methods can be purely data-driven, more mechanistic models develop explicit cause-and-effect hypotheses about biological or epidemiological drivers of transmission dynamics. Previous work has compared performance between these approaches, finding no evidence of substantial variation in predictive accuracy between these model structures (4–6).

An alternative way of looking at model development is to consider a model’s specificity towards the target setting to which the forecasts are being applied. One approach to model building is to create a model able to produce forecasts for any given setting, given only updated data. This is the case, for example, for typical time-series forecasting and most semi-mechanistic models. A different approach is to adapt the model structure itself to the predictive target, for example to emphasise different mechanistic processes or to exploit differences in the availability and quality of multiple data sources. Such context-specific models may be more policy-relevant (7,8), and a study of COVID-19 forecasts in Germany and Poland suggested the two best performing models were those most tuned to the national context (9).

However, it has been difficult to establish clear associations between different choices in model development, such as model structure and specificity, and the predictive performance of resulting forecasts. Challenges include a lack of comparability between forecasts (10), a small sample size of participating modelling teams (11), and the need to account for varying predictive difficulty among multiple forecast targets with differing outbreak characteristics (12).

A number of collaborative epidemic forecasting efforts have aimed to support such analyses. Such “Hubs” collate predictions from diverse participating modellers, often publishing these projections in real-time during an epidemic. Alongside informing public health, this enables like-for-like comparisons of predictive performance (13). While there may be inherent limits to predictability (14), several factors have been suggested to contribute to general predictive ability. For example, infectious disease dynamics may be easier to predict in the short term (15), when they are more stable (9), among larger population sizes (16), reported with timely surveillance (6), or have leading indicators in complementary data sources (17).

Analyses of forecast performance have typically used a univariate approach to describing differences in accuracy between models. However, such findings could be better supported by using more formal models of association that are also able to capture factors affecting overall predictability (11,16,18). Here, we used a general additive model with forecast scores as an outcome while including covariate indicators of predictive difficulty. We evaluate associations between model structure and geographic specificity with predictive performance, in the context of short-term forecasts of COVID-19 in Europe.

## Methods

We evaluated forecasts from the European COVID-19 Forecast Hub between 8 March 2021 to 10 March 2023. This is a public platform launched in March 2021 that collated real-time weekly forecasts of COVID-19 for 32 European countries (19). Over 50 modelling teams contributed prospective forecasts during the study period, predicting between one and four weeks ahead incidence of COVID-19 cases, deaths, or hospitalisations. Forecasters were able to express uncertainty by reporting a distribution of up to 23 given probabilistic quantiles for each prediction. Forecasters also contributed a standard set of metadata describing their team and model. Contributions were voluntary, and the number and identity of forecasters contributing to the Hub project varied over time.

We assessed forecasts of incident cases or deaths and excluded targets for hospitalisations, which experienced multiple changes in source data during the study period. We excluded forecasts that did not report the full set of 23 quantiles, and those that did not forecast over the entire four week horizon, in order to ensure fair comparison among probabilistic model results (see Supplement). We also excluded baseline and ensemble models created by the Hub team from evaluation.

We categorised each model in terms of model structure using the metadata provided by each modelling team, according to the following types (in order of decreasing mechanistic detail): “Agent-based” (with detailed individual-level interactions), “Mechanistic” (broadly based on compartmental models), “Semi-mechanistic” (statistical models with epidemiologically informed elements, e.g. delay distributions and/or generation intervals) and “Statistical” (context-agnostic time-series models). We further classified models as “Other” that were ensembles of human-judgement based predictions. We based this on teams’ short description of their methods included in metadata, and any additional links provided by teams, such as citations or websites with model details. Four members of the research team (KS, SF, FB, JM) were tasked with classifying models, with at least three investigators independently classifying each model. The majority of choices determined the final classification and any potential ties flagged for an additional independent classification by another investigator. Two investigators (SF, KS) separately reviewed all classifications in discussion for any final revisions.

We also classified models by geographic target specificity. We assessed the total number of countries that each model had provided a projection for at any time over the forecast period, noting that forecasters could change the number of targets they submitted. We classified each model as “single” or “multi-country” based on forecasting for one versus two or more countries.

We compared forecasts against data collated by Johns Hopkins University (JHU) comprising cumulative reported cases or deaths reported daily for each of the 32 European countries (20). This had been communicated to participating teams as being the “truth” data set. We calculated the incident weekly number of cases or deaths and used the dataset available as of 10 March 2023, when JHU stopped reporting new data. We excluded forecasts after this time in order to use a single consistent data source against which to evaluate forecasts.

We evaluated forecasts using the Weighted Interval Score (WIS). This assigns forecasts a score as a combination of dispersion, overprediction, and underprediction, reflecting both the width of the forecast and whether it was centred above or below the observed value (21). We log-transformed all forecasts and observations before applying the weighted interval score, which leads to more comparable scores across locations and times, and can be interpreted as scoring the ability of models to predict the growth rate (22,23).

We used summary statistics to describe the performance as measured by WIS in our dataset by model structure and geographic target specificity. In order to make more robust inferences about effects on forecast performance, we further fitted a generalised additive mixed effect model (GAMM). We used the WIS of each individual forecast as an outcome variable (separately for cases and deaths) and used a Gaussian error model with a log-link (see Supplement). We added a small amount (1e-7, or less than 1/1000th of the smallest positive value in the data) to values in order to avoid zeros. Explanatory variables included model structure, and whether a model targeted one or multiple countries, each modelled with random effects.

We adjusted for five factors that we believed could confound associations with forecast performance, selected a priori. First, forecast performance typically declines with longer forecast horizons, and we included length of horizon in weeks as a factor smooth spline (separately for each forecasting model). Second, we also considered the epidemic trend in the target week, using observed data to retrospectively categorise each week as “Stable”, “Decreasing”, or “Increasing”, based on the difference over a three-week moving average of incidence (with a change of +/-5% as “Stable”; Supplement Figs 2 & 3). We included this as a random effect. In order to account for any differences in difficulty of forecasting different locations/times, we included the specific geographic target as a random effect; and a smooth spline per country target over time. Last, we added a random effect for individual models to account for any variation in accuracy beyond the other factors included in our model.

In order to demonstrate the impact of adjusting for confounding factors, we further fitted univariate models for each of the explanatory variables and by individual model, and contrasted unadjusted and adjusted estimates of the partial effect of each factor. We fitted models with the *mgcv* package in R (24), using restricted maximum likelihood estimation (REML). All data and code are publicly available online at (25) (with details in Supplement).

## Results

We evaluated a total 181,851 forecast predictions from 47 forecasting models, contributed by 37 separate modelling teams to the European COVID-19 Forecast Hub (Table 1). 5 teams contributed more than one model. Participating models varied over time as forecasting teams joined or left the Hub and contributed predictions for varying combinations of forecast targets. Between 7 and 33 models contributed in any one week, forecasting for any combination of 256 possible weekly forecast targets (32 countries, 4 horizons, and 2 target outcomes). On average each model contributed 3869 forecasts, with the median model contributing 764 forecasts.

**Table 1:** Characteristics of forecasts sampled from the European COVID-19 Forecast Hub, March 2021-2023. Forecast performance was measured using the weighted interval score (WIS), with a lower score indicating a more accurate forecast.

| Variable | Cases |  |  | Deaths |  |  |
| --- | --- | --- | --- | --- | --- | --- |
|  | Models | Forecasts | Mean WIS (SD) | Models | Forecasts | Mean WIS (SD) |
| Overall | 42 (100%) | 91966 (100%) | 0.57 (0.93) | 38 (100%) | 89885 (100%) | 0.37 (0.51) |
| **Method** |  |  |  |  |  |  |
| Agent-based | 3 (7.1%) | 814 (0.9%) | 0.44 (0.58) | 2 (5.3%) | 720 (0.8%) | 0.25 (0.25) |
| Mechanistic | 16 (38.1%) | 27987 (30.4%) | 0.54 (0.88) | 16 (42.1%) | 33449 (37.2%) | 0.36 (0.46) |
| Semi-mechanistic | 9 (21.4%) | 28742 (31.3%) | 0.59 (1) | 10 (26.3%) | 28494 (31.7%) | 0.38 (0.6) |
| Statistical | 11 (26.2%) | 32680 (35.5%) | 0.58 (0.95) | 7 (18.4%) | 25478 (28.3%) | 0.39 (0.46) |
| Other | 3 (7.1%) | 1743 (1.9%) | 0.33 (0.43) | 3 (7.9%) | 1744 (1.9%) | 0.29 (0.41) |
| **Number of country targets** |  |  |  |  |  |  |
| Single-country | 19 (45.2%) | 3680 (4%) | 0.41 (0.49) | 14 (36.8%) | 2821 (3.1%) | 0.28 (0.32) |
| Multi-country | 23 (54.8%) | 88286 (96%) | 0.57 (0.95) | 24 (63.2%) | 87064 (96.9%) | 0.38 (0.51) |

We categorised 12 models as statistical, 12 as semi-mechanistic, 17 as mechanistic, 3 as agent-based and 3 models that used human judgement forecasting as “other” (Supplementary Table 1). For 17 (36%) models, investigators disagreed on model classification. The majority of 2/3 was used as the final classification, with additional manual review which in all cases retained the majority decision. In the volume of forecasts provided, mechanistic, semi-mechanistic, and statistical models each contributed similar numbers of forecasts with approximately one-third each. Agent-based and “other” models provided fewer forecasts, representing only 1-2% of forecasts.

We considered models forecasting for only one, or multiple countries. We collated 19 single-country models and 28 multi-country models. Single-country models targeted Germany (7 models), Poland (5), Spain (3), Italy (2), Czechia (2) and Slovenia (1). The average multi-country model forecast for a median number of 23 locations. Models classified as targeting multiple countries could vary from week to week in how many locations they forecast. Only 2 models consistently forecast for the same number of locations throughout the entire study period.

We explored the interval score (WIS) as a measure of predictive performance (Fig 1), and characterised its association with model structure and number of countries targeted. We used descriptive statistics and an unadjusted univariate model for each explanatory variable, and then a generalised additive mixed model to give adjusted estimates of the partial effect of each factor while controlling for other sources of variation (Fig 2). The interval score was highly right-skewed with respect to all explanatory variables, which we accounted for by using a log-link.

**Figure 1.**
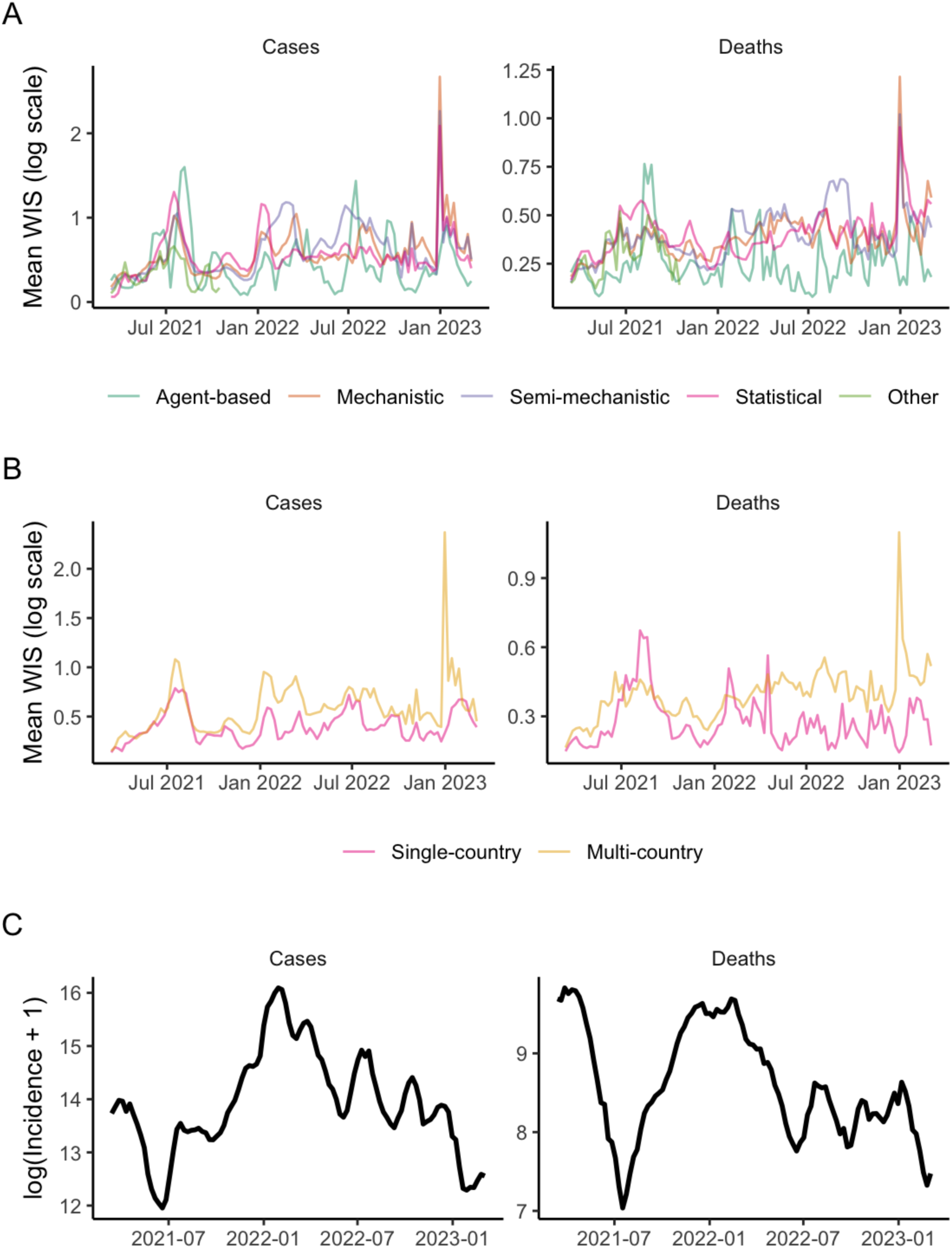
Predictive accuracy of multiple models’ forecasts for COVID-19 cases and deaths across 32 European countries over time. Forecast performance is shown as the mean weighted interval score (WIS), where a lower score indicates better performance. Forecast performance is summarised across 32 target locations and 1 through 4 week forecast horizons, with varying numbers of forecasters participating over time. Shown for (A) the method structure used by each model; (B) the number of countries each model targeted (one or multiple); with (C) the total count of observed incidence across all 32 countries, shown on the log scale.

**Figure 2.**
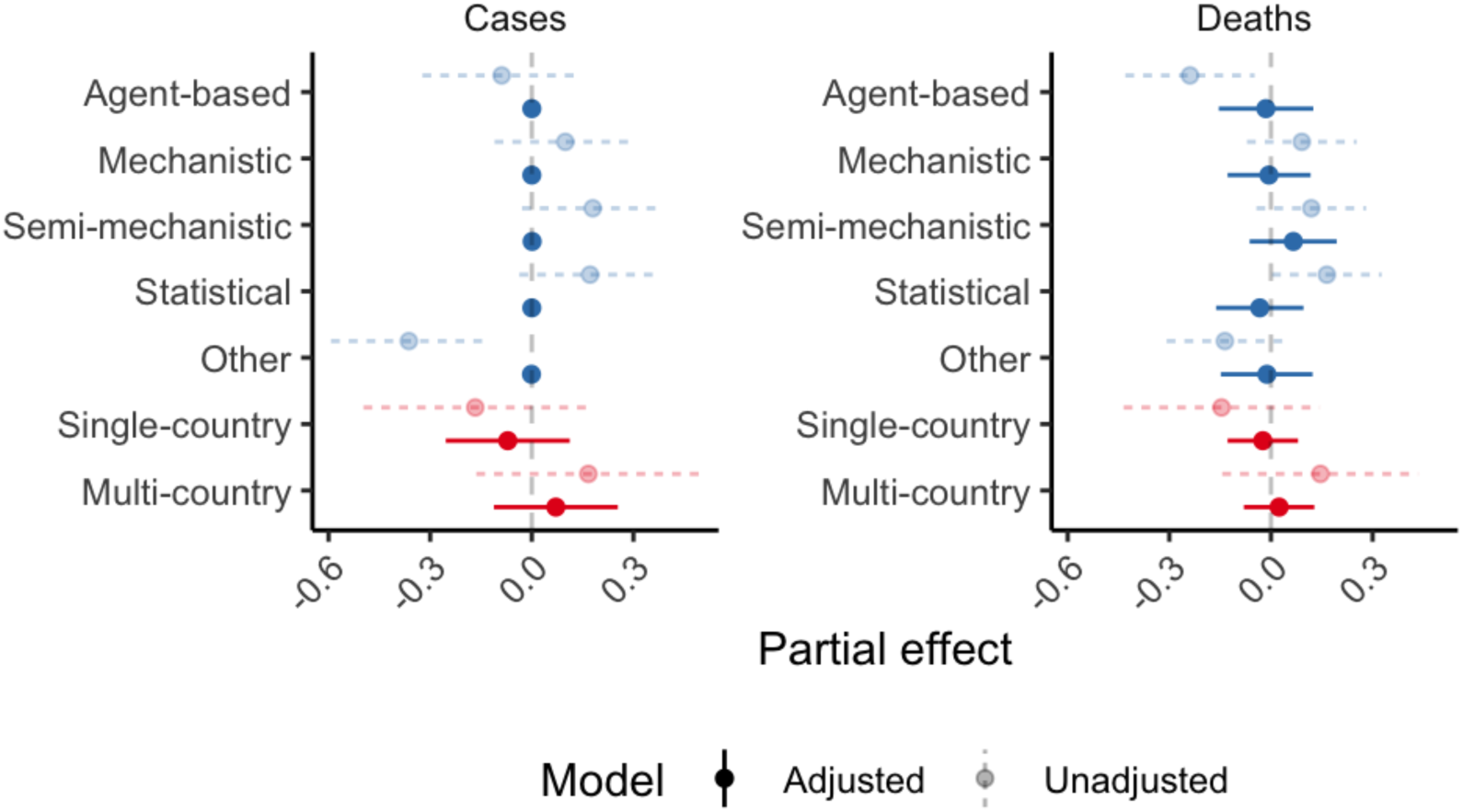
Effect on the weighted interval score (WIS) of model structure and number of countries targeted, before and after adjusting for confounding factors. A lower WIS indicates better performance, meaning effects <0 are relatively better than the group average. Adjusted effects also account for the impact of forecast horizon, epidemic trend, geographic location, and individual model variation. Partial effects and 95% confidence intervals were estimated from fitting a generalised additive mixed model.

Descriptively, we noted apparently similar predictive performance between mechanistic, semi-mechanistic, and statistical models. These model structures appeared to perform relatively worse than agent-based and “other” models. For example, in univariate analysis, the partial effect for statistical models forecasting deaths indicates underperformance by 0.165 (95%CI 0.003-0.327) compared to average, while agent-based models performed better than average (-0.24 (-0.43– 0.05)). However, variation in performance overlapped between all model structures, and we noted relative differences between models may have varied over time (Fig 1). For example, over summer 2021 all model types saw worsening performance coinciding with the introduction of the Delta variant across Europe, but this decline was most marked among statistical models of death outcomes compared to any other model type.

These differences between model structures largely disappeared after adjustment for our five covariates (forecast horizon, epidemic trend, country, country over time, and individual model). We found no clear evidence that any one type of model consistently outperformed others (Fig 2). There was no difference in accuracy between model structures when predicting cases, and we observed only weak differences when predicting deaths. In contrast to unadjusted estimates, we identified that statistical models may have performed slightly better (partial effect-0.03 (-0.16-0.1)), and semi-mechanistic models worse (0.07 (-0.06-0.19)) than the average, although with overlapping uncertainty.

Considering the number of countries targeted by each model, we descriptively noted that single-country models typically out-performed compared to multi-country models. This relative performance was stable over time, although with overlapping ranges of variation. Multi-country models appeared to have a more sustained period of poorer performance in forecasting deaths from spring 2022, although we did not observe this difference among case forecasts. We noted multi-country models may have had particular difficulty forecasting around January 2023, both with respect to cases and deaths.

In adjusted estimates, we also saw some indication that models focusing on a single country outperformed those modelling multiple countries (partial effect for single-country models forecasting cases:-0.07 (-0.25-0.11), compared to 0.07 (-0.11-0.25) for multi-country models; and-0.02 (-0.13-0.08) and 0.02 (-0.08-0.13) respectively when forecasting deaths). However, these effects were inconclusive with overlapping uncertainty.

We considered the predictive horizon and epidemiological situation for each forecast as potentially confounding other associations with model performance. In unadjusted estimates, average performance worsened from 1 to 4 weeks of predictive horizon, and was best when the epidemiological situation was stable. Model based analysis gave weak support to this observation. This indicated improved predictive performance during stable periods of each country’s outbreak curve (cases:-0.24 (-0.57-0.08); deaths:-0.19 (-0.41-0.04)), compared to increasing trends in incidence (cases:-0.07 (-0.39-0.25); deaths:-0.02 (-0.24-0.2)), with worst performance seen when predicting decreasing trends (cases: 0.31 (-0.01-0.64); deaths: 0.2 (-0.02-0.43)).

We identified residual unexplained influences among models’ performance. We interpreted the estimated partial random effect for each model as a proxy of its performance whilst correcting for missingness and the common factors considered here. We noted substantial variation beyond the factors we included, indicating that our variable selection was not sufficient for fully explaining performance (Fig 3).

**Figure 3.**
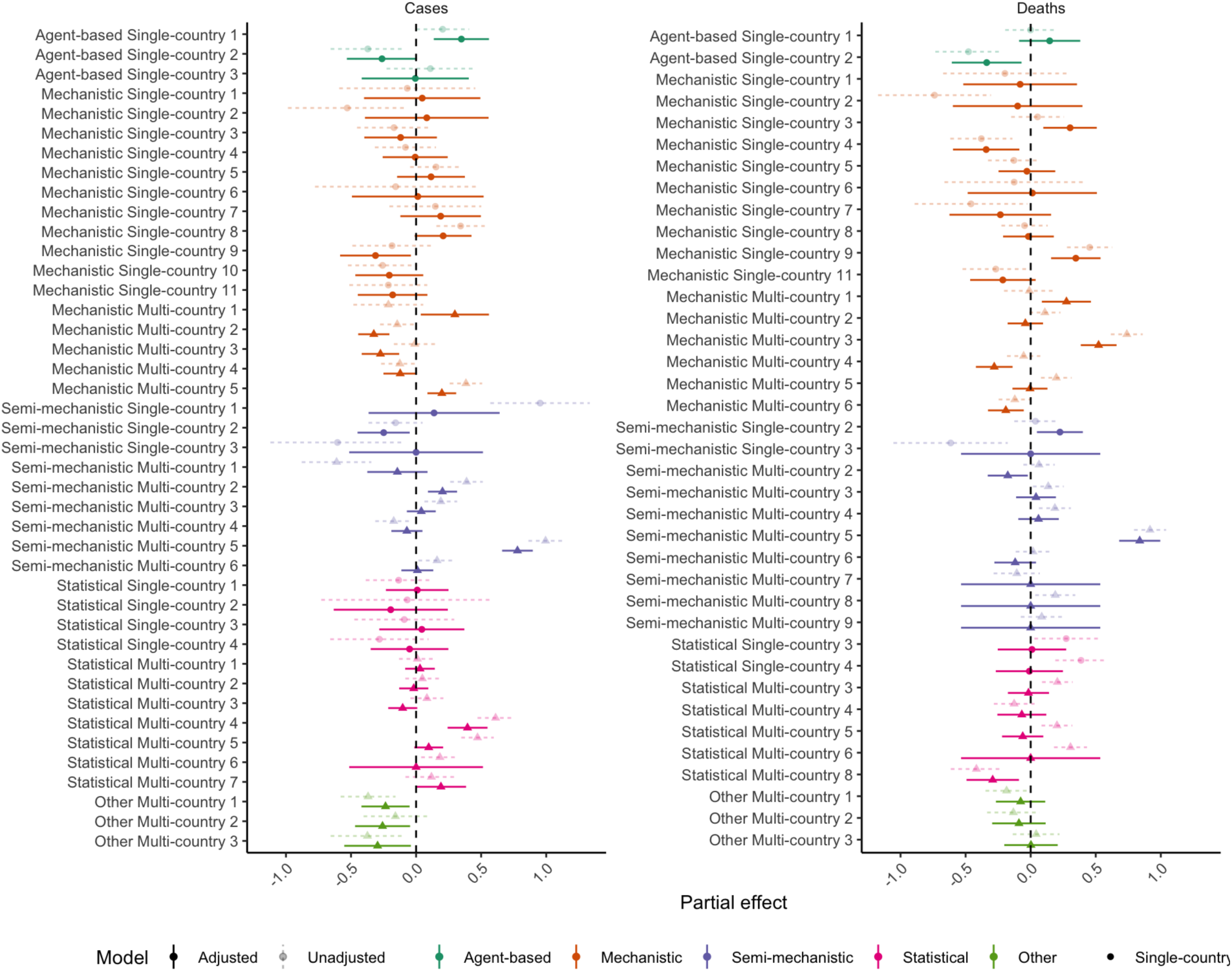
Partial effect size (95% CI) by model. This can be interpreted as adjusted performance after accounting for all other variables in the model, with remaining differences in effects as seen here representing unexplained variation between models beyond these factors.

## Discussion

We aimed to assess influences on the accuracy of infectious disease forecasts, considering models’ methodological structure and the specificity of the model to the geographic target. Using data from the European COVID-19 Hub, we assessed comparable forecasts across 32 countries with independent epidemic processes, evaluating 181,851 predictions from 47 forecasting models. We used a mixed effects model to identify forecast performance while accounting for confounding factors affecting general predictive difficulty. After adjustment, we observed little systematic difference in average performance between model structures. Models forecasting for only a single geographic target outperformed those that made predictions for multiple targets, although this was a weak signal with overlapping uncertainty. We noted substantial variation in model performance that our approach did not account for.

Our findings extend previous work that has failed to find evidence for substantial differences between different model types’ predictive performance, across a variety of outbreaks and epidemic dynamics (4–6). In contrast, some prior moderate evidence suggests that statistical methods gave better performance than more mechanistic approaches (17,18,26). The typical advantage of statistical models is that they capture the autocorrelated nature of incidence time series. However, we also noted that the more mechanistic models in our study produced a wide range of predictive performance, and some of the best-performing individual models in our sample were mechanistic or agent-based. This indicates that factors beyond our structural categorisation may have played a role in driving performance. At the same time, we note widely replicated work demonstrating that the most reliable method for achieving predictive accuracy is to combine forecasts across models and model types using ensemble methods, rather than relying on any single model (e.g. (12,15,27)).

We observed some weak evidence that models tended to perform better when they were focused on single countries versus multiple countries. This could suggest that the flexibility to incorporate knowledge about a specific forecast target can yield a performance advantage. This could derive from a better understanding of the nature of the underlying data generating processes that modellers were asked to forecast, such as country-specific policies influencing local epidemic dynamics, or the specific way in which the public data reflected underlying transmission processes. Other work has similarly noted forecaster experience as a contributing factor in improving performance (18).

We also noted that real-time data were frequently updated retrospectively, and such data revisions might also explain some of the improved performance of target-specific models. Teams focussing on only one country’s data may have been more easily able to detect anomalous observations or be aware of their cause, and consequently to adapt forecasts to predict a value closer to the “true” observation that would later be reflected in a retrospective data revision (e.g. (28)).

A key limitation of this work was its limited power to detect potential differences in performance between model structures or target specificity. While we used a large dataset of forecasts over time, we had a relatively small sample size of independent models: for example, only 3 agent-based models. Further, the Hub relied on an opportunistic cohort of models from voluntary contributions. This meant our results could be biased by unobserved characteristics that differentially affected participation, such as the resource intensity of model development. A larger or more systematic sample of models would support a deeper investigation of factors driving model performance. Future work could approach this by combining data from across multiple similar Hub projects and a more detailed assessment of the characteristics of the model cohort.

Our model-based approach to comparing forecast performance while controlling for variability across targets could be improved and developed further. Firstly, there are likely many interactions between the effects explored here. For example, a modeller’s choice of model structure may interact with their chosen number of targets, as in agent-based models where all three represented here targeted a single country. In particular, different model structures may be more or less well equipped to predict at each interval of the one to four week forecast horizon. For example, models incorporating underlying epidemic mechanisms may be less responsive to shorter term data artefacts but relatively better able to demonstrate the impact of longer term dynamics. Explicitly accounting for such interactions could be a useful insight from future work.

Further, we did not account for a wide range of additional confounding factors likely influencing performance. These may include the presence of anomalies in real-time data, or varying uses of model assumptions, or data sources (11). We did not attempt to quantify the amount of resources or effort that teams put into model development or producing forecasts, or explore how this might relate to model structures that require more or less tuning and input. In addition, we did not track changes to each model’s individual methods over time, or account for this in our analysis. Further work could particularly investigate in more detail the interactions between methodological choices and the broader context of epidemiological predictability, for example accounting for turning points or changes to underlying transmission dynamics such as new variant introductions or management interventions.

Unlike past work, our conclusions are based on assessing performance using forecasts on a log scale, rather than the natural scale of observed data. We observed that the log scale moderated scores overall, and in general this method appears to give less weight to penalising inaccurate forecasts after a peak (23). In particular, we saw reduced variability among the scores of semi-mechanistic models on the log scale, compared to the natural scale. This suggests the importance of accounting for epidemic characteristics in assessing forecast performance, where we chose to prioritise relative assessment of the growth rate rather than absolute counts. We think that interpreting performance against trends in underlying epidemic dynamics better reflects decision maker needs. It remains important to bear in mind that our conclusions towards the chosen metric may not hold when choosing other scoring rules. We also note that we modelled the interval score using a Gaussian error model with a log-link. This impacted the propriety of the score as effects were multiplicative, making estimates unsuitable for direct comparison or ranking performance among individual models.

In this study we contributed to better understanding the drivers of predictive performance among forecasting models, and highlighted the importance of appropriately adjusting for the general predictive difficulty of the forecast target. Comparisons like the one we have conducted here support a more informed view of the role of modelling in pandemic preparedness and response. Further model comparison projects should enable this by encouraging methodological diversity among participants, and ensuring that detailed structured information on model metadata (such as methodology and model revisions) is collated alongside numerical predictions. We would particularly encourage studies that can assess the amount of local context that modellers use in creating or adapting their forecast model, and similarly whether performance scales with efforts to continually revise models, incorporate domain knowledge, or evaluate model performance in the real-time forecasting process. Future performance evaluations could combine information from multiple collaborative efforts and ultimately lead to a convergence of recommendations for which models to use when making epidemiological forecasts in an epidemic or pandemic.

## Data Availability

The codebase for this paper is publicly available at the Github repository “epiforecasts/eval-by-method” and Zenodo with DOI: 10.5281/zenodo.14903162. Forecast and observed data were sourced from the European COVID-19 Forecast Hub, available to view online. All Hub data are now archived at the Github repository: “european-modelling-hubs/covid19-forecast-hub-europe_archive” and Zenodo with DOI: 10.5281/zenodo.13986751. Specific data used in this work are available in the Github repository for this paper.

https://github.com/epiforecasts/eval-by-method

https://covid19forecasthub.eu/

https://github.com/european-modelling-hubs/covid19-forecast-hub-europe_archive

https://doi.org/10.5281/zenodo.13986751

## Supporting Information

**Supplementary Figure 1**. Eligibility criteria for models contributing case and death forecasts to the European COVID-19 Forecast Hub, March 2021 - March 2023

**Supplementary Table 1.** Model characteristics contributing to the European COVID-19 Forecast Hub, by method used, number of countries targeted, and number of forecasts contributed

**Supplementary Figure 2**. Epidemic trends (cases)

**Supplementary Figure 3**. Epidemic trends (deaths)

**Supplementary Figure 4**. Model diagnostics (cases)

**Supplementary Figure 5**. Model diagnostics (deaths)

## Notes

### Competing Interest Statement

The authors have declared no competing interest.

### Funding Statement

KS, SF, SA were funded by Wellcome Trust (grant number 200901/Z/16/Z).

### Author Declarations

Source data were openly available before the initiation of the study. Forecast and observed data were sourced from the European COVID-19 Forecast Hub, available to view at ˂https://covid19forecasthub.eu/˃. All Hub data are now archived at Github: ˂https://github.com/european-modelling-hubs/covid19-forecast-hub-europe_archive˃ and Zenodo with DOI: ˂https://doi.org/10.5281/zenodo.13986751˃. Specific data used in this work are available in the Github repository for this paper at: ˂https://github.com/epiforecasts/eval-by-method/tree/main/data˃

