## Supplementary Information for "The influence of model structure and geographic specificity on predictive accuracy among European COVID-19 forecasts"

### 1 Code and data availability

#### 1.1 Code

The codebase for this paper is publicly available at:

- Github: https://github.com/epiforecasts/eval-by-method
- Zenodo with DOI: https://doi.org/10.5281/zenodo.14903162

Comments and code contributions are welcome - please use Github Issues.

Please cite code using:

- Katharine Sherratt & Sebastian Funk. (2025). epiforecasts/eval-by-method: Zenodo. https://doi.org/10.5281/zenodo.14903162

#### 1.2 Source data

Forecast and observed data were sourced from the European COVID-19 Forecast Hub, available to view at https://covid19forecasthub.eu/ . All Hub data are now archived at:

- Github: https://github.com/european-modelling-hubs/covid19-forecast-hub-europe_archive
- Zenodo with DOI: https://doi.org/10.5281/zenodo.13986751

Data for this work were downloaded on 30th May 2023. These data are available in the Github repository for this paper at: https://github.com/epiforecasts/eval-by-method/tree/main/data

#### 2 Model characteristics

##### 2.1 Eligibility criteria

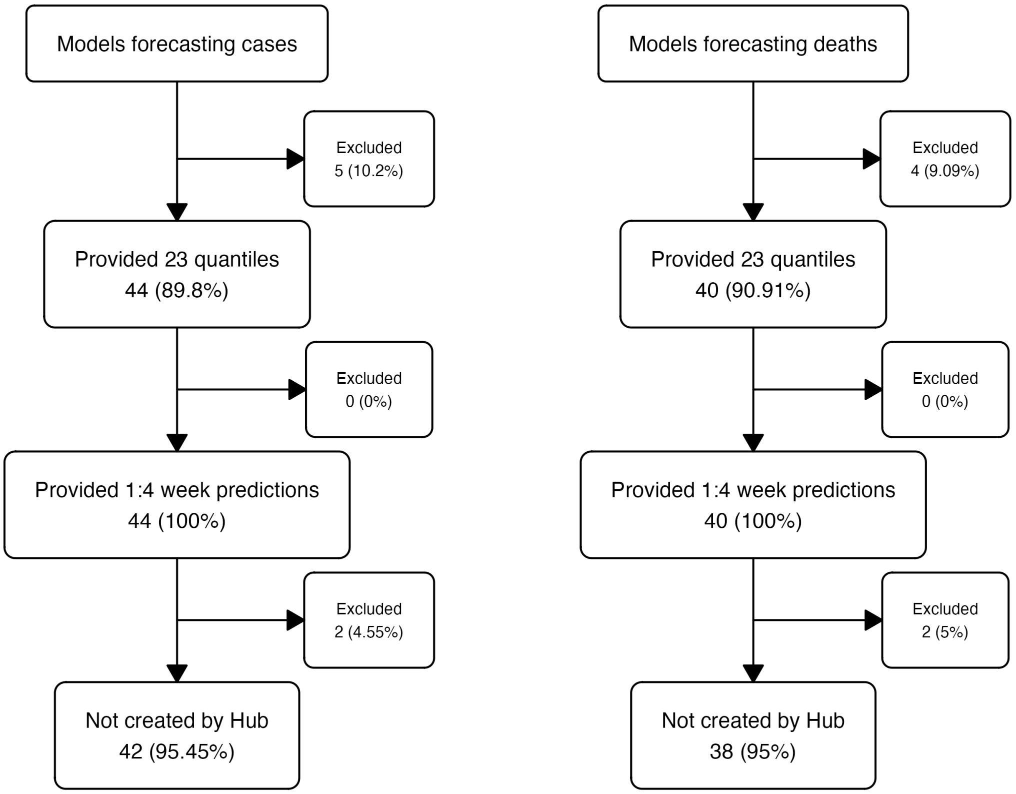

Figure 1: Eligibility criteria for models contributing case (left) and death (right) forecasts to the European COVID-19 Forecast Hub, March 2021 - March 2023

##### 2.2 Model characteristics

Table 1: Model characteristics contributing to the European COVID-19 Forecast Hub, by method used, number of countries targeted, and number of forecasts contributed.

| Model | Method | Country Targets | Case forecasts | Death forecasts |
|---|---|---|---|---|
| AMM-EpiInvert | Statistical | Multi-country | 2,788 (1.3%) | |
| CovidMetrics-epiBATS | Statistical | Single-country | 343 (0.2%) | |
| DSMPG-bayes | Semi-mechanistic | Multi-country | 760 (0.4%) | |
| EuroCOVIDhub-baseline | Statistical | Multi-country | 13,082 (6.3%) | 13,040 (6.3%) |
| FIAS_FZJ-Epi1Ger | Mechanistic | Single-country | 264 (0.1%) | 264 (0.1%) |
| GoeWroc-BaseBayes | Semi-mechanistic | Single-country | 12 (0%) | |
| HZI-AgeExtendedSEIR | Mechanistic | Single-country | 382 (0.2%) | 382 (0.2%) |
| ICM-agentModel | Agent-based | Single-country | 334 (0.2%) | 334 (0.2%) |
| IEM_Health-CovidProject | Mechanistic | Multi-country | 7,710 (3.7%) | 7,708 (3.7%) |
| ILM-EKF | Semi-mechanistic | Multi-country | 11,998 (5.8%) | 11,961 (5.8%) |
| ITWW-county_repro | Semi-mechanistic | Single-country | 650 (0.3%) | 600 (0.3%) |
| Imperial-DeCa | Semi-mechanistic | Multi-country | | 571 (0.3%) |
| Imperial-RtI0 | Semi-mechanistic | Multi-country | | 571 (0.3%) |
| Imperial-sbkp | Semi-mechanistic | Multi-country | | 571 (0.3%) |
| JBUD-HMXK | Mechanistic | Multi-country | 1,324 (0.6%) | 1,324 (0.6%) |
| KITmetricslab-bivar_branching | Statistical | Single-country | 8 (0%) | |
| Karlen-pypm | Mechanistic | Multi-country | 3,208 (1.5%) | 3,186 (1.5%) |
| LANL-GrowthRate | Semi-mechanistic | Multi-country | 3,692 (1.8%) | 3,696 (1.8%) |
| LeipzigIMISE-SECIR | Mechanistic | Single-country | 16 (0%) | 16 (0%) |
| MIMUW-StochSEIR | Mechanistic | Single-country | 76 (0%) | 76 (0%) |
| MIT_CovidAnalytics-DELPHI | Mechanistic | Single-country | 348 (0.2%) | 500 (0.2%) |
| MOCOS-agent1 | Agent-based | Single-country | 386 (0.2%) | 386 (0.2%) |
| MUNI-ARIMA | Statistical | Multi-country | 10,979 (5.3%) | 11,314 (5.4%) |
| MUNI-LaggedRegARIMA | Statistical | Multi-country | | 736 (0.4%) |
| MUNI-VAR | Statistical | Multi-country | 976 (0.5%) | 976 (0.5%) |
| MUNI_DMS-SEIAR | Mechanistic | Single-country | 224 (0.1%) | 200 (0.1%) |
| PL_GRedlarski-DistrictsSum | Mechanistic | Single-country | 378 (0.2%) | |
| RobertWalraven-ESG | Statistical | Multi-country | 9,190 (4.4%) | 10,465 (5%) |
| SDSC_ISG-TrendModel | Statistical | Multi-country | 1,756 (0.8%) | 1,744 (0.8%) |
| UB-BSLCoV | Statistical | Single-country | 96 (0%) | 96 (0%) |
| UC3M-EpiGraph | Agent-based | Single-country | 94 (0%) | |
| ULZF-SEIRC19SI | Mechanistic | Single-country | 249 (0.1%) | 249 (0.1%) |
| UMass-MechBayes | Mechanistic | Multi-country | | 5,948 (2.9%) |
| UMass-SemiMech | Semi-mechanistic | Multi-country | 1,888 (0.9%) | 1,904 (0.9%) |
| UNED-PreCoV2 | Statistical | Single-country | 147 (0.1%) | 147 (0.1%) |
| UNIPV-BayesINGARCHX | Statistical | Multi-country | 426 (0.2%) | |
| USC-SIkJalpha | Mechanistic | Multi-country | 12,900 (6.2%) | 12,688 (6.1%) |
| UpgUmibUsi-MultiBayes | Semi-mechanistic | Single-country | 99 (0%) | 99 (0%) |
| bisop-seirfilter | Mechanistic | Single-country | 32 (0%) | 32 (0%) |
| bisop-seirfilterlite | Mechanistic | Multi-country | 336 (0.2%) | 336 (0.2%) |
| epiMOX-SUIHTER | Mechanistic | Single-country | 134 (0.1%) | 134 (0.1%) |
| epiforecasts-EpiExpert | Other | Multi-country | 945 (0.5%) | 948 (0.5%) |
| epiforecasts-EpiExpert_Rt | Other | Multi-country | 404 (0.2%) | 404 (0.2%) |
| epiforecasts-EpiExpert_direct | Other | Multi-country | 394 (0.2%) | 392 (0.2%) |
| epiforecasts-EpiNow2 | Semi-mechanistic | Multi-country | 8,843 (4.3%) | 7,721 (3.7%) |
| epiforecasts-weeklygrowth | Statistical | Multi-country | 5,971 (2.9%) | |
| itwm-dSEIR | Mechanistic | Single-country | 406 (0.2%) | 406 (0.2%) |
| prolix-euclidean | Semi-mechanistic | Multi-country | 800 (0.4%) | 800 (0.4%) |

### 3 Statistical methods

#### 3.1 Epidemic trend identification

We retrospectively categorised each week as “Stable”, “Decreasing”, or “Increasing”, based on the difference over a three-week moving average of incidence (with a change of +/-5% as “Stable”).

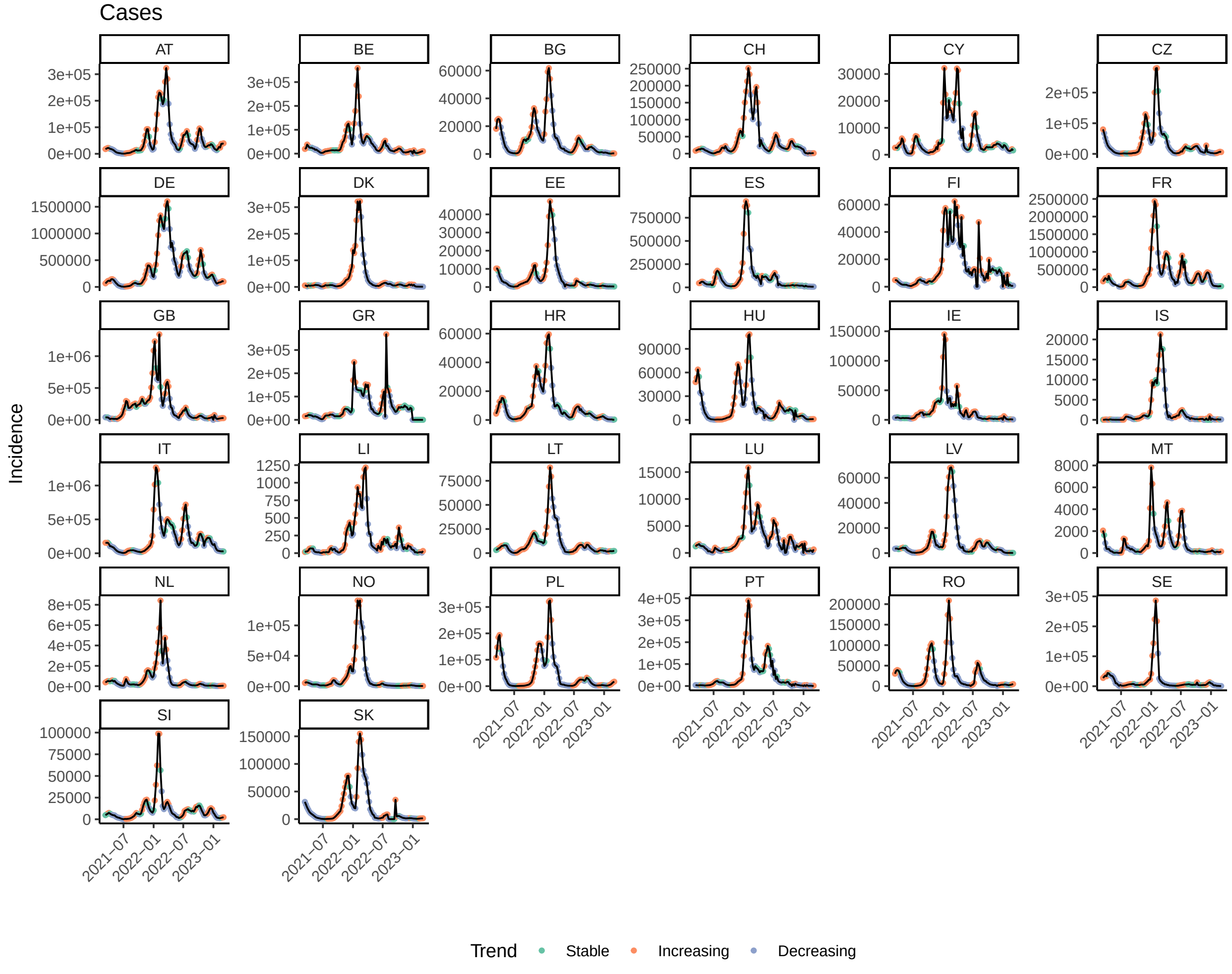

Figure 2: Trends (cases)

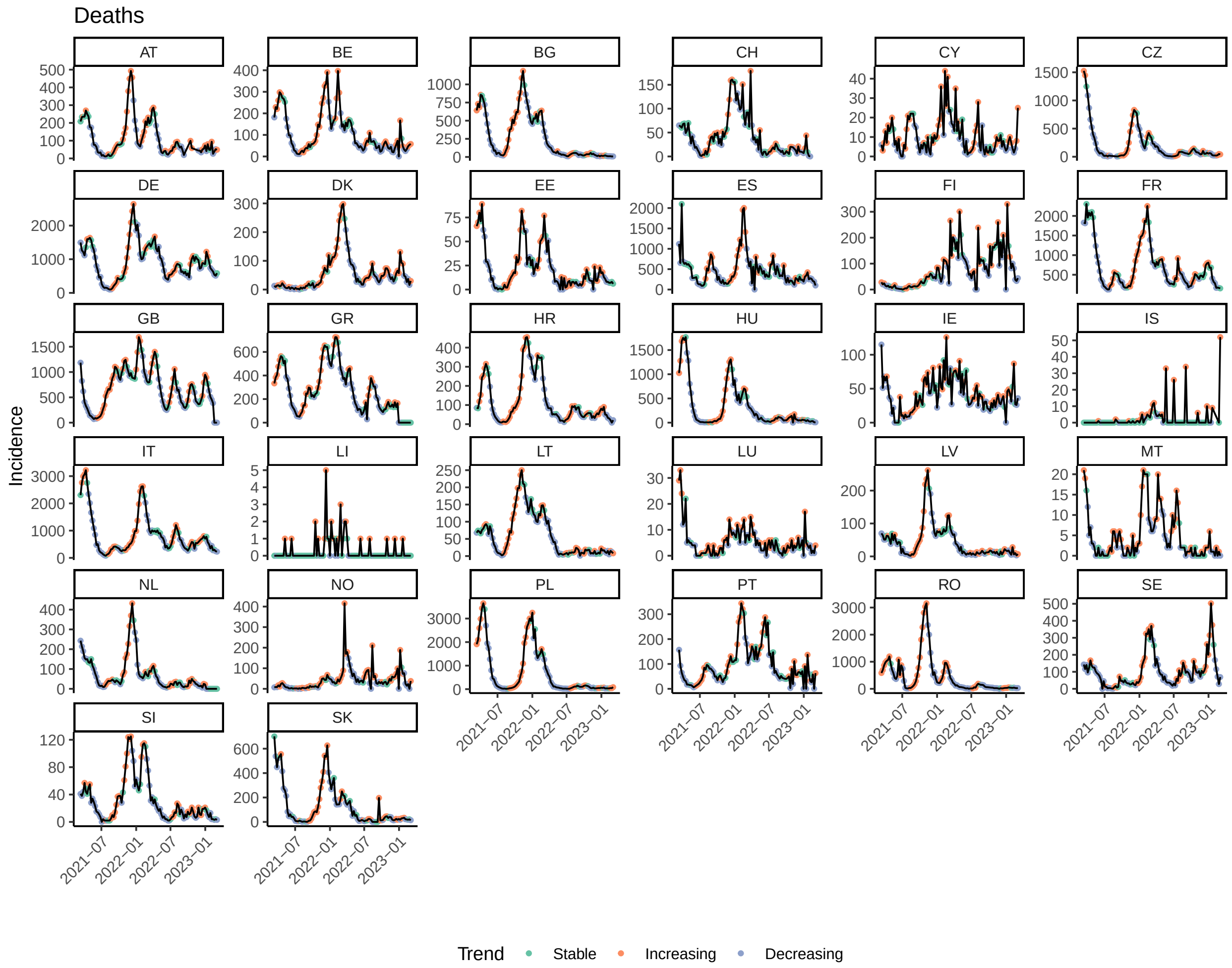

Figure 3: Trends (deaths)

#### 3.2 Model fitting

#### 3.3 Model formula

$\sim$, wis, s(Method, bs = "re") + s(CountryTargets, bs = "re") + s(Trend, bs = "re") + s(location, bs = "re") + s(time, by = location, k = 40) + s(Horizon, k = 3, by = Model, bs = "sz") + s(Model, bs = "re")

#### 3.4 Model diagnostics

##### 3.4.1 Cases

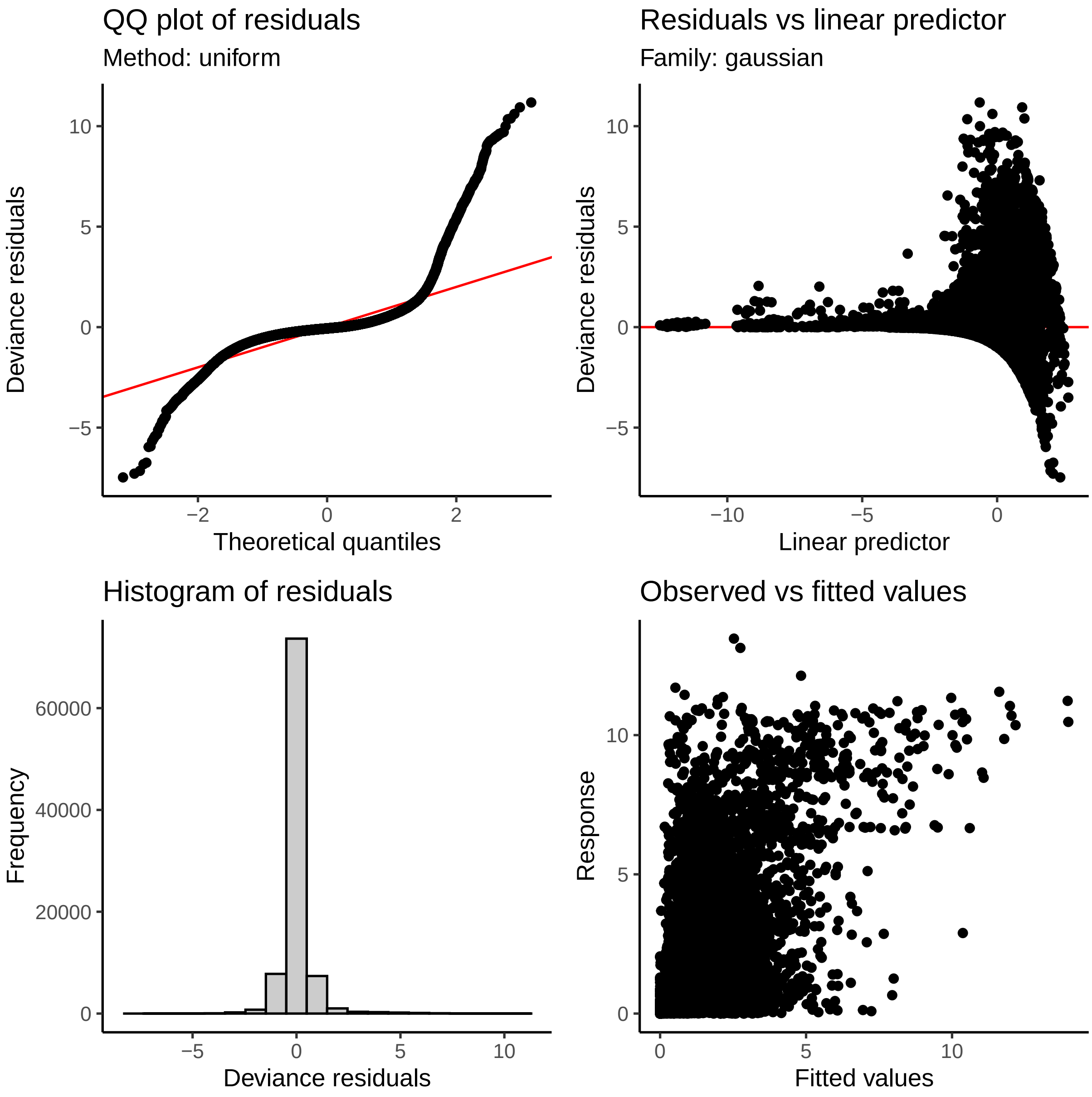

##### 3.4.2 Deaths

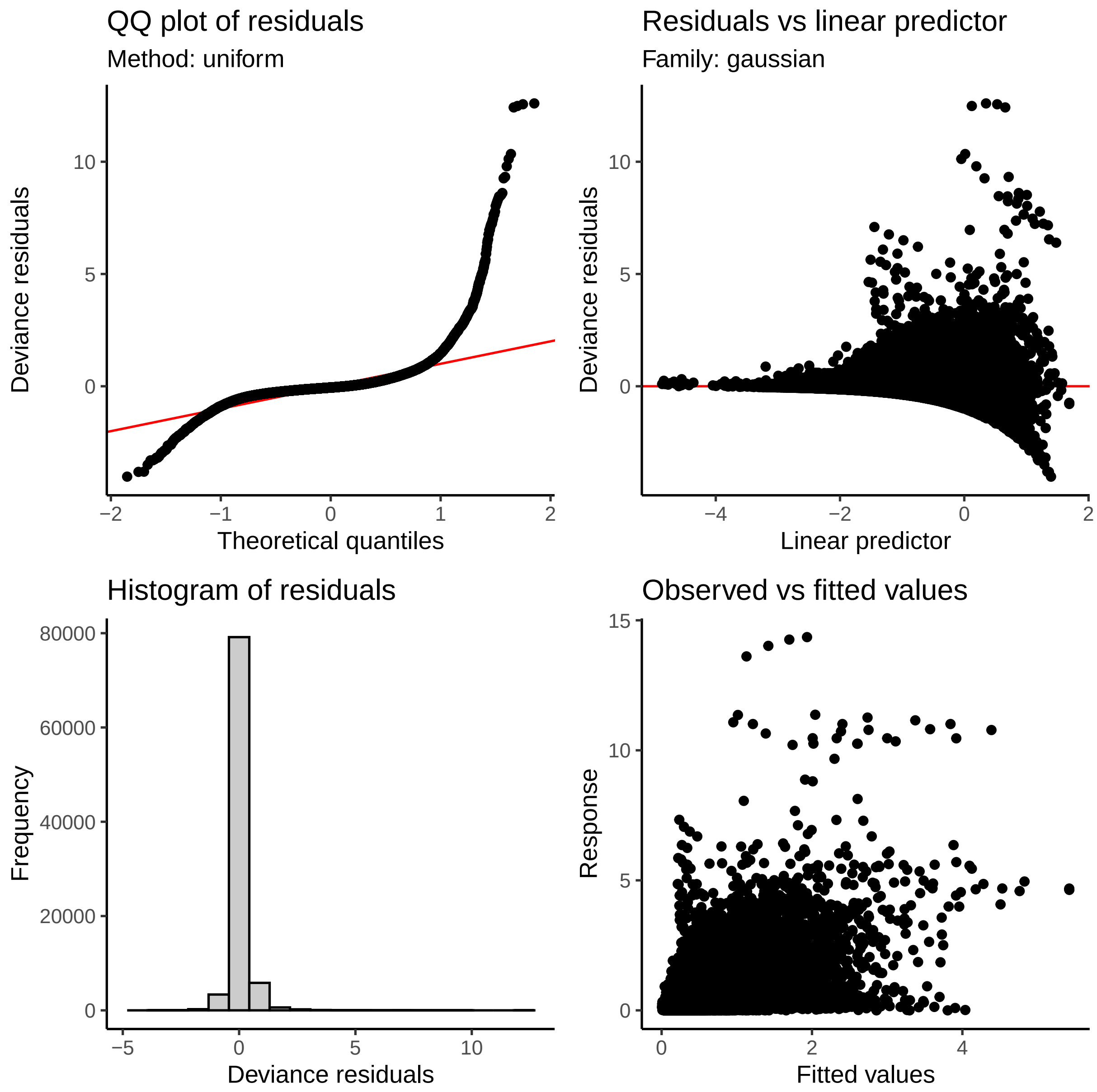
QQ plot of residuals
Method: uniform
Deviance residuals
Theoretical quantiles
Residuals vs linear predictor
Family: gaussian
Deviance residuals
Linear predictor
Histogram of residuals
Frequency
Deviance residuals
Observed vs fitted values
Response
Fitted values
